# Factors Influencing the Use of Multiple HIV Prevention Services among Men in the General Population of Kenya: An analysis of the 2022 KDHS

**DOI:** 10.1101/2025.07.04.25330883

**Authors:** John Baptist Asiimwe, Lilian Nuwabaine, Benjamin Betunga, Quraish Sserwanja, Joseph Kawuki

## Abstract

**Background:** The use of multiple HIV prevention strategies is more effective in preventing HIV than single-method approaches. This study, therefore, aimed to examine the factors influencing the utilization of multiple HIV prevention methods among Kenyan men.

**Methods:** Secondary data from the 2022 Kenya Demographic and Health Survey (KDHS) involving 14,453 participants selected by multistage stratified sampling were analysed. Multivariable logistic regression was conducted using SPSS (version 29) to identify associated factors.

**Results:** About 75.4% of men used at least two HIV prevention methods. Most were circumcised (94.2%), followed by HIV testing (73.5%) and condom use at last sexual intercourse (30.2%). Concerning the use of multiple HIV prevention services, the findings indicated that a relatively low proportion of the men who had tested for HIV, were circumcised and then used condoms (22.7%), followed by men who had tested for HIV, and then used condoms (23.9%), and those who were circumcised and then used condoms (28.7%). Factors associated with utilization of multiple HIV prevention services included HIV knowledge (aOR=4.64 (95%CI: 1.34-16.11), age at first sex (aOR=0.25 (95%CI: 0.08-0.79), age at first child (aOR=10.37 (95%CI: 1.26-85.60), number of lifetime partners (aOR=0.44 (95%CI: 0.24-0.80), access to newspapers (aOR=2.77 (95%CI: 1.42-5.43)), tribe (Taita/Taveta= aOR=0.02 (95%CI: 0.003-0.16), Meru (aOR=0.20 (95%CI: 0.06-0.70), and Luo= aOR=0.05 (95%CI: 0.01-0.18)) and justified refusing of sex by their partners (aOR=2.03 (95%CI:1.12-3.66), North Easten religion (aOR=0.10(95%CI:0.02-0.56), Eastern (aOR=0.23(95%CI:0.07-0.77), Central (aOR=0.16(95%CI:0.04-0.62), Rift valley (aOR=0.06(95%CI:0.02.0.18)

**Conclusion:** Utilization of multiple HIV prevention services among Kenyan men is above average, but sociodemographic and community factors influenced the use of multiple HIV prevention services among men. Innovative targeted awareness campaigns about the HIV risk and benefits of multiple HIV prevention services are needed for older men, those from certain tribes, regions, and with multiple sex partners. However, stakeholders need to involve and incorporate cultural and religious leaders to raise awareness about the benefits of prevention services and ensure their accessibility. There is a need to incorporate gender-transformative messaging in HIV outreach programs and mass media to promote HIV service use among men. The government of Kenya should implement region-specific interventions, including mobile clinics and culturally sensitive health promotion, to strengthen community health systems in underserved regions.

## Introduction

Human Immunodeficiency Virus (HIV) remains a significant public health challenge worldwide, with sub-Saharan Africa (SSA) bearing a disproportionate share of the burden (1). As of 2021, Sub-Saharan Africa accounted for approximately 67% of all people living with HIV globally (2). In Kenya, the adult HIV prevalence was estimated at 4.9% in 2017, with approximately 1.5 million people living with HIV. Notably, the prevalence among men aged 15 to 49 years was 3.5%, compared to 6.2% among women in the same age group (3). Comparatively, some countries in Western Africa and Southern Africa exhibited lower HIV prevalence rates. For instance, Nigeria and Zambia had adult HIV prevalence rates between 2-3% (4). In contrast, regions such as Western and Central Europe and North America reported lower HIV prevalence rates, often below 1% (1). This disparity underscores the unique challenges faced by countries like Kenya in combating the HIV epidemic (4).

Despite not meeting the global target of less than 370,000 new HIV infections by 2025, there has been a decrease in the HIV incidence by 39% reduction since 2010, reducing from 2.1 million to 1.3 million in 2023 (5, 6). However, certain regions have experienced increases in new infections. For example, in East Africa, there has been a rise in HIV incidence rates, attributed to factors such as intravenous drug use and limited access to prevention services. In Kenya, it was estimated that approximately 22,000 new HIV infections occurred in 2022, translating to about 60 new infections daily (7).

Addressing the HIV epidemic among men in Kenya is crucial for several reasons. Men often have lower rates of HIV testing and are less likely to engage in care compared with women, leading to delayed diagnosis and treatment (8). This delay increases the risk of HIV transmission and contributes to higher mortality rates (8). Failure to tackle HIV among men undermines global initiatives such as Sustainable Development Goal (SDG) 3, which aims to ensure healthy lives and promote well-being for all ages, including ending the AIDS epidemic by 2030 (7).

Using multiple HIV prevention strategies is more effective than single-method approaches (5). For instance, the combination of condom use and voluntary medical male circumcision (VMMC) was found to significantly reduce the risk of HIV acquisition (9). In Kenya, offering multiple HIV prevention services to key populations led to a 44% decrease in new HIV infections, while in Vietnam, Cambodia, and Thailand, new infections dropped by more than 60% from 2010 to 2020 (6).

Despite the proven benefits, the uptake of multiple HIV prevention services among men in Kenya remains largely unknown (10). However, a study among transport workers in southwestern Uganda found that less than half (45.3%) of participants had used HIV testing services and condoms within one year (11). The Kenyan government has implemented various initiatives to promote the use of multiple HIV prevention services. These include the distribution of free condoms, scaling up VMMC programs, and increasing access to HIV testing and counselling services (3). However, gaps persist, particularly among men. Stigma, discrimination, and legal barriers hinder the effective utilization of these services among men (12).

Several factors that influence the utilization of multiple HIV prevention services among men at risk for HIV in Uganda and other countries have been identified. These factors included HIV knowledge, education, perceived HIV risk, stigma and discrimination, accessibility and availability of HIV services (11). Men in rural areas or regions with limited healthcare infrastructure may have reduced access to multiple prevention services (12). While these factors have been identified in various studies, they have focused on the single use of HIV prevention services among risky groups, ignoring the general population where most men belong. Therefore, there is a paucity of research specifically examining the determinants of multiple HIV prevention service utilization among men who do not fall under risk groups in Kenya using nationally representative data. Understanding these factors is essential to inform targeted interventions and policies aimed at increasing the uptake of multiple HIV prevention services among men in Kenya. Therefore, this study aimed to explore the factors influencing the utilization of multiple HIV prevention methods among men in Kenya.

## Methods

### Data source, sample design, and collection

This research used the 2022 Kenya Demographic and Health Survey (KDHS), which implemented a two-stage stratified sampling technique. In the initial stage, 1692 enumeration areas (EAs) or clusters were selected from a master sample frame consisting of 129,067 EAs derived from the 2019 Kenya population and housing census, using equal probability with independent selection (3). This was followed by a house listing to establish a sampling frame, which was then used in the second stage to select 25 households from each cluster. Nonetheless, if a cluster contained fewer than 25 households, all of them were included in the sample. Ultimately, the survey was conducted across 1,691 clusters. The Inner-City Fund (ICF) supervised the pretesting of the research instruments and trained the data collection team; data was gathered from February to July 2022. Interviews were conducted in Swahili or English with all men aged 15-54 years who were regular members of the chosen households or had stayed overnight in those households the night before the survey. (3). From a total of 14,818 men who participated in the survey, 14,453 men were considered for this analysis, resulting in a response rate of 98% (3). Despite the dataset having hundreds of variables, we only took into account those that were pertinent and applicable to our research.

### Study variables Dependent/outcome

Considering the data available in KDHS, the main focus of this study was the utilization of multiple HIV prevention services.a composite variable constructed from 3 binary (yes/no) variables, namely whether one had: 1) been circumcised or not at the time of the interview, 2) used a condom at their last sex, and 3) ever tested for HIV (3). Using two or more of the above-mentioned services (≥2) was considered as using multiple HIV prevention services, a binary outcome variable (Yes/No). Although antiretroviral drug therapy use was not included in the analysis of multiple HIV services use, findings for those who had been diagnosed with HIV in the general population are reported.

### Independent variables

The factors included in the analysis were categorized into four, namely, sociodemographic, lifestyle behavioural, sexual behavioural, psychosocial factors, and intimate partner violence factors (11, 13). Thirteen sociodemographic factors were included in the analysis, namely; household size (≥5 vs ≤4 members), age in years (35–54, 25–34, 15–24), ethnicity (categorized into the twelve tribes of Kenya; Taita/Taveta, Embu, Mijikenda/Swahili, Kalenjin, Maasai, Kikuyu, Kamba, Luhya, Kisii, Luo, Meru, Somali, and Others), education (tertiary, secondary, none/primary), region (classified into Kenya’s eight provinces; Northeastern, Eastern, Coast, Western, Nairobi, Central, Rift Valley, anc Nyanza), residence (urban vs rural), marital status (unmarried vs married/cohabiting), working status (No vs Yes), religion (Christian, Muslim, or others), and wealth index (richest, poorer, richer, poorest and middle). The wealth index was derived from household asset ownership data using principal component analysis (3). The participants’ perceived health status at the time of interview (bad, moderate, good), and exposure to mass media such as internet, radio, television, and newspapers (No vs Yes) were also included in the analysis. Autonomy was assessed through four proxy variables, namely, who makes decisions related to healthcare-seeking, earnings, major household purchases, (jointly with partner/other person, partner, self, or others) and who heads the household (female vs male).

Three lifestyle behavioural factors included whether the participants and their sex partners (wives) consumed alcohol or used tobacco (yes/no). We also examined ten variables that were related to sexual behaviours which included the age of the participant at first sexual intercourse in years (≤14, 15-24, ≥25), early fatherhood as indicated by the age of respondent at their first childbirth (≤14, 15-24, ≥25), recent sexual activity (active vs inactive), number of sex partners in last 12 months (1 vs ≥2), number of women the participant had children with as a proxy indicator for engaging in a polygamous relationship (1 or ≥2), number of living children (≤2, 3-4, ≥5), number of lifetime sex partners (≤5, 6-10, >10), HIV serostatus (negative vs positive), and any symptoms of sexually transmitted infections (STIs) in the last 12 months preceding the survey in the form of a genital ulcer or discharge (yes/no).

Four psychosocial factors included the desire for more children (wants, does not want, undecided), whether their wives were justified to refuse sex if they had other women (yes/no), whether their wives were justified to ask their husbands to use condoms if he has STI (yes/no), HIV knowledge and stigma. Knowledge of HIV was assessed as a binary variable based on 5 questions regarding HIV transmission and prevention, with response options being “No”, “Yes”, or “Don’t Know”. A correct response for each item was awarded a score of 1, with a maximum score of 5, which was further categorised as poor (≤2) or good knowledge (≥3).

Whereas HIV stigma was computed from two items measuring stigmatizing attitudes towards people living with HIV (yes/no). Agreeing to both or any of the two statements indicated that one possessed HIV stigma (yes) and vice versa if one agreed to none of the statements (No).

Intimate partner violence (IPV) related factors were also included in the analysis, namely whether the participant was afraid of their wife/partner (yes/no), justified beating (yes/no), and experiencing controlling, emotional, physical, and sexual forms of IPV (yes/no). It is worth noting that justified beating (5 items), controlling (5 items), emotional (4 items), physical (7 items) and sexual (3 items) IPV variables were composite variables constructed from numerous binary response items, where agreement to any of the items (≥1) indicated justified beating or an experience of any forms of IPV.

### Statistical analysis

The data were analyzed using the SPSS (version 29) complex samples package, which took into consideration the complex sample design characteristic of DHS data that involves stratification and clustering during participant sampling (14, 15). Prior to the analysis, the dataset was cleaned, and dummy variables were created. At the univariate level, descriptive statistics, including frequencies, were calculated for all categorical variables. However, to address the unequal probability sampling across various strata and to guarantee the study results’ representativeness, DHS sample weights were utilized for all computed frequencies (14, 15). Univariate and multivariate logistic regression were used to obtain independent factors associated with the use of multiple HIV services. All variables with P-values <0.1 were entered in simple multivariate logistic regression to establish the factors associated with the use of multiple HIV services. All variables’ odds ratios are reported at 95% confidence intervals. Multi-collinearity was also assessed among all the predictor variables in the model using a variance inflation factor (VIF) of greater than 10 as a cutoff (14, 15). None of the factors exceeded the cutoff.

### Ethical consideration

No ethical clearance was required to analyze the secondary data since it is publicly accessible. However, authorization to access the 2022 KDHS datasets was secured from MEASURE DHS (https://www.dhsprogram.com/data/available-datasets.cfm). Ethical clearance for conducting the study, documented in the datasets, was granted by the Institutional Review Board of the Inner-City Fund (ICF). The study was carried out by the Kenya National Bureau of Statistics in partnership with various stakeholders. Written informed consent was obtained from both human subjects and legally designated representatives of minor participants.

### Characteristics of the study participants

A total of 14,453 men participated in this study (see Table 1). The majority of the participants resided in rural areas (61.1%), were between the ages of 15 and 34 years (66.7%), and identified as being from the Eastern, Central, Rift Valley, and Nairobi regions (67.3%). Most identified as belonging to the Kalenjin, Luhya, Kikuyu, and Kamba ethnic groups (58.3%), were Christians (85.5%), employed (78.2%), unmarried (51.9%), and had completed at least secondary education (79.6%). In total, 65.8% belonged to the middle, richer, and richest quintiles, 95% had more than five household members, and 82.9% lived in male-headed households. Most participants made decisions to seek healthcare services on their own (55.9%); however, decisions regarding household purchases (53.7%) and earnings (50.1%) were made jointly with their partners. Most men had access to mass media, which encompassed television (80.1%), radio (87.4%), the Internet (58.8%), and newspapers (39.4%), and reported that they were in good health (84.1%). Regarding lifestyle habits, 26.6% of men consumed alcohol, while 13.3% engaged in tobacco use, and 5.1% had wives/partners who consumed alcohol. The majority were recently sexually active (51.2%), with no permanent sex partners in the last 12 months (66.4%), but had started sex at the age of 15-24 (72.2%) and had ≤ 5 lifetime sexual partners (64.3%). Many of the men had two children (≤ 2, 71.9%), had their first child at the age of 15-24years (45.7%), most had those children with one woman/sex partner (79.6%), and desired to have more children (53.6%). Although a few men had STI symptoms in the form of a genital discharge or ulcer (2.1%), the majority were HIV negative (98.8%). Most participating men had good knowledge of HIV (96.2%), no stigmatizing attitudes (80.7%), and believed that women (their wives) were justified in refusing sex if they had other women (79.0%) or asking their husbands to use condoms if they had STIs (90.7%). Few participants were afraid of their wives (7.7%) and justified that their wives/sex partners should be beaten for numerous reasons (27.0%). The majority of men had experienced IPV from their wives/ sex partners, mostly in the form of controlling (74.0%), emotional (71.9%), physical (7.5 %), and sexual violence (5.0%).

**Table 1.** Characteristics of the study participants.

| **Variable (N=14453)** | **n (weighted% )** |
| --- | --- |
| **Sociodemographic factors** |  |
| **Age (years)** |  |
| 15-24 | 5579 (38.6) |
| 25-34 | 4055 (28.1) |
| 35-54 | 4819 (33.3) |
| **Residence** |  |
| Urban | 5624 (38.9) |
| Rural | 8829 (61.1) |
| **Region/Province** |  |
| Coast | 1341 (9.3) |
| Northeastern | 273 (1.9) |
| Eastern | 2123 (14.7) |
| Central | 1945 (13.5) |
| Rift Valley | 3808 (26.3) |
| Western | 1499 (10.4) |
| Nyanza | 1610 (11.1) |
| Nairobi | 1855 (12.8) |
| **Ethnicity** |  |
| Mijikenda/Swahili | 798 (5.5) |
| Kalenjin | 1823 (12.6) |
| Kamba | 1669 (11.5) |
| Kikuyu | 2521 (17.4) |
| Kisii | 901 (6.2) |
| Luhya | 2424 (16.8) |
| Luo | 1591 (11.0) |
| Maasai | 325 (2.2) |
| Meru | 717 (5.0) |
| Embu | 186 (1.3) |
| Taita/Taveta | 136 (0.9) |
| Somali | 294 (2) |
| Other | 1068 (7.4) |
| **Education** |  |
| None/primary | 5660 (39.1) |
| Secondary | 5851(40.5) |
| Tertiary | 2943 (20.4) |
| **Religion** |  |
| Christians | 12361 (85.5) |
| Muslims | 1042 (7.2) |
| Others | 1050 (7.3) |
| **Marital status** |  |
| Married/Cohabiting | 6958 (48.1) |
| Unmarried (single, divorced/widowed/separated) | 7495 (51.9) |
| **Working status** |  |
| Working | 11296 (78.2) |
| Not working | 3157 (21.8) |
| **Wealth index** |  |
| Poorest | 2175 (15.1) |
| Poorer | 2762 (19.1) |
| Middle | 2932 (20.3) |
| Richer | 3504 (24.2) |
| Richest | 3079 (21.3) |
| **Sex of Household head** |  |
| Male | 11978 (82.9) |
| Female | 2475 (17.1) |
| **Household number** |  |
| ≤4 | 717 (5) |
| ≥5 | 13736 (95) |
| **Health status** |  |
| Bad | 189 (1.3) |
| Moderate | 2116 (14.6) |
| Good | 12148 (84.1) |
| **Health-seeking decision-making** |  |
| Self | 3886 (55.9) |
| Joint | 2644 (38.0) |
| Partner | 399 (5.7) |
| Others | 29 (0.4) |
| **Purchase decision making** |  |
| Self | 2323 (33.4) |
| Joint | 3738 (53.7) |
| Partner | 883(12.7) |
| Others | 14 (0.2) |
| **Earnings decision making** |  |
| Self | 2777 (42.6) |
| Joint | 3270 (50.1) |
| Partner | 473 (7.3) |
| Others | 1 (0.01) |
| **Newspaper** |  |
| Yes | 5700 (39.4) |
| No | 8753 (60.6) |
| **TV** |  |
| No | 2875 (19.9) |
| Yes | 11578 (80.1) |
| **Radio** |  |
| Yes | 12633 (87.4) |
| No | 1820 (12.6) |
| **Internet use** |  |
| Yes | 8496 (58.8) |
| No | 5957 (41.2) |
| **Lifestyle Behavioural factors** |  |
| **Tobacco use** |  |
| Yes | 1928 (13.3) |
| No | 12525 (86.7) |
| **Alcohol consumption** |  |
| Yes | 3845 (26.6) |
| No | 10608 (73.4) |
| **Wife drinks Alcohol** |  |
| Yes | 243 (5.1) |
| No | 4484 (94.9) |
| **Sexual Behavioural Factors** |  |
| **Age at first sex (years)** |  |
| 14 | 2812 (23.1) |
| 15-24 | 8791 (72.2) |
| ≥25 | 574 (4.7) |
| **Sexual activity** |  |
| Active | 7396 (51.2) |
| Inactive | 7057 (48.8) |
| **Number of sex partners in the last 12 months** |  |
| None | 7495 (66.4) |
| 1 partner | 6613 (24.6) |
| ≥2 partners | 345 (8.9) |
| **Number of Women with respondents' children** |  |
| 1 partner | 6194 (79.6) |
| ≥2 partners | 1585 (20.4) |
| **Number of lifetime sex partners** |  |
| ≤5 | 7826 (64.3) |
| 6-10 | 2260 (18.6) |
| >10 | 2091 (17.2) |
| **HIV status** |  |
| Negative | 10415 (98.8) |
| Positive | 126 (1.2) |
| **Age of respondent at 1st childbirth** |  |
| 14 | 62 (0.8) |
| 15-24 | 3556 (45.7) |
| ≥25 | 4161 (53.5) |
| **Genital ulcer in the last 12 months** |  |
| Yes | 304 (2.1) |
| No | 14135 (97.9) |
| **Genital Discharge in the last 12 months** |  |
| Yes | 297 (2.1) |
| No | 14120 (97.9) |
| **Number of living children** |  |
| ≤2 | 10385 (71.9) |
| 3-4 | 2520 (17.4) |
| ≥5 | 1548 (10.7) |
| **Psychosocial factors** |  |
| **Desire for more children** |  |
| Wants | 3717 (53.6) |
| Does not want | 2816 (40.6) |
| Undecided | 401 (5.8) |
| **HIV knowledge** |  |
| Poor | 369 (3.8) |
| Good | 9221 (96.2) |
| **Stigma** |  |
| Yes | 2781 (19.3) |
| No | 11620 (80.7) |
| **Wife justified to ask husband to use a condom** |  |
| Yes | 12873 (90.7) |
| No | 1325 (9.3) |
| **Wife justified in refusing sex with husband.** |  |
| Yes | 11143 (79.0) |
| No | 2970 (21.0) |
| **IPV factors** |  |
| **Respondent is afraid of their wife/partner** |  |
| Yes | 364 (7.7) |
| No | 4363 (92.3) |
| **Justified beating** |  |
| Yes | 3808 (27.0) |
| No | 10284 (73.0) |
| **IPV** |  |
| Yes | 3500 (74.0) |
| No | 1228 (26.0) |
| **Emotional IPV** |  |
| Yes | 1203 (25.5) |
| No | 3524 (74.5) |
| **Controlling IPV** |  |
| Yes | 3397 (71.9) |
| No | 1330 (28.1) |
| **Physical IPV** |  |
| Yes | 355 (7.5) |
| No | 4372 (92.5) |
| **Sexual IPV** |  |
| Yes | 237 (5.0) |
| No | 4490 (95.0) |

HIV: Human Immunodeficiency Virus, IPV =intimate partner violence

### The use of multiple HIV prevention services

Overall, although 98% (95%CI:97.7-98.3) of the men in Kenya had used at least one of the four HIV prevention services evaluated in this study, only 75.4% (95%CI:74.3-76.4) had ever used two or more services (≥2) or multiple HIV prevention methods (**Table 2**). The largest percentage of men were circumcised (94.2% (95%CI:93.6-94.8) followed by those who used anti-retroviral therapy among the HIV positive participants (90.9% (95%CI:83.2-95.3), HIV testing services (73.5% (95%CI:72.4-74.6), and condoms at last sexual intercourse (30.2% (95%CI:28.6-31.8). Concerning the use of multiple HIV prevention services, the findings indicated that a relatively low proportion of the men had tested for HIV, were circumcised and then used condoms (22.7% (95%CI: 21.4-24.1), followed by men who had tested for HIV, and then used condoms (23.9% (95%CI: 22.5-25.5), and those who were circumcised and then used condoms (28.7% (95%CI: 27.3-30.2).

**Table 2.** Use of HIV prevention services among men in Kenya

| Variable | N | Weighed% (95%CI) |
| --- | --- | --- |
| **Overall (at least one method)** | **14453** | **98 (97.7-98.3)** |
| **Use of multiple HIV services (2 or more methods) \*** | **14453** | **75.4 (74.3-76.4)** |
| Male circumcision | 14443 | 94.2 (93.6- 94.8) |
| Condom use at last sex | 10773 | 30.2 (28.6- 31.8) |
| Antiretroviral therapy | 126 | 90.9 (83.2- 95.3) |
| HIV testing | 14453 | 73.5 (72.4- 74.6) |
| HCT+ condom use | 10773 | 23.9 (22.5-25.5) |
| SMC+ condom use | 10769 | 28.7 (27.3-30.2) |
| HCT+SMC+condom use | 10769 | 22.7 (21.4- 24.1) |

CI: confidence interval. HCT=HIV counselling and Testing, SMC=Male circumcision.
\*=Given the small sample size for those taking ART (n=126), we did not include the ART HIV service in the computation for the use of multiple HIV services, as it may provide misleading results.

### Factors associated with the use of multiple HIV prevention services

The factors associated with using multiple HIV prevention methods in the univariate and multivariate logistic regression analysis are summarized in **Table 3**. We found that compared with those with poor knowledge, men who had good knowledge of HIV were 4.64 (95%CI: 1.34-16.11) times more likely to use multiple HIV prevention methods. Participants with access to newspapers were more likely to use multiple HIV prevention methods when compared to those without access (aOR=2.77, 95%CI: 1.42-5.43). Similarly, participants who believed that their wives were justified in refusing sex if their husbands had other women were 2.03 (95%CI: 1.12-3.66) times more likely to use multiple HIV prevention methods than those who thought to the contrary. Additionally, compared with those who had their first sex at a younger age (15-24 years), men who had their first sex at an older age (≥25 years) were less likely to use multiple HIV prevention methods (aOR=0.25, 95%CI: 0.08-0.79). However, men who had their first child at an older age (≥15 years) compared with the younger (≤15 years) were more likely to use multiple HIV prevention methods (e.g., ≥25 years= aOR 10.37 (95%CI: 1.26-85.60). In addition, compared with men who had ≤5 lifetime partners, men who had more than 6-10 lifetime sexual partners were 0.44 (95%CI: 0.24-0.80) times less likely to use multiple HIV prevention methods. Similarly, when compared with those from the Kikuyu tribe, participating men from certain tribes of Kenya had lower odds of using multiple HIV prevention methods (Taita/Taveta= aOR=0.02 (95%CI: 0.003-0.16), Meru (aOR=0.20 (95%CI: 0.06-0.70), and Luo= aOR=0.05 (95%CI: 0.01-0.18). Likewise, men from some provinces of Kenya were less likely to use multiple HIV prevention methods (e.g., Northern eastern with aOR=0.10, 95%CI: 0.02-0.56) when compared with those from the coastal region.

**Table 3.**
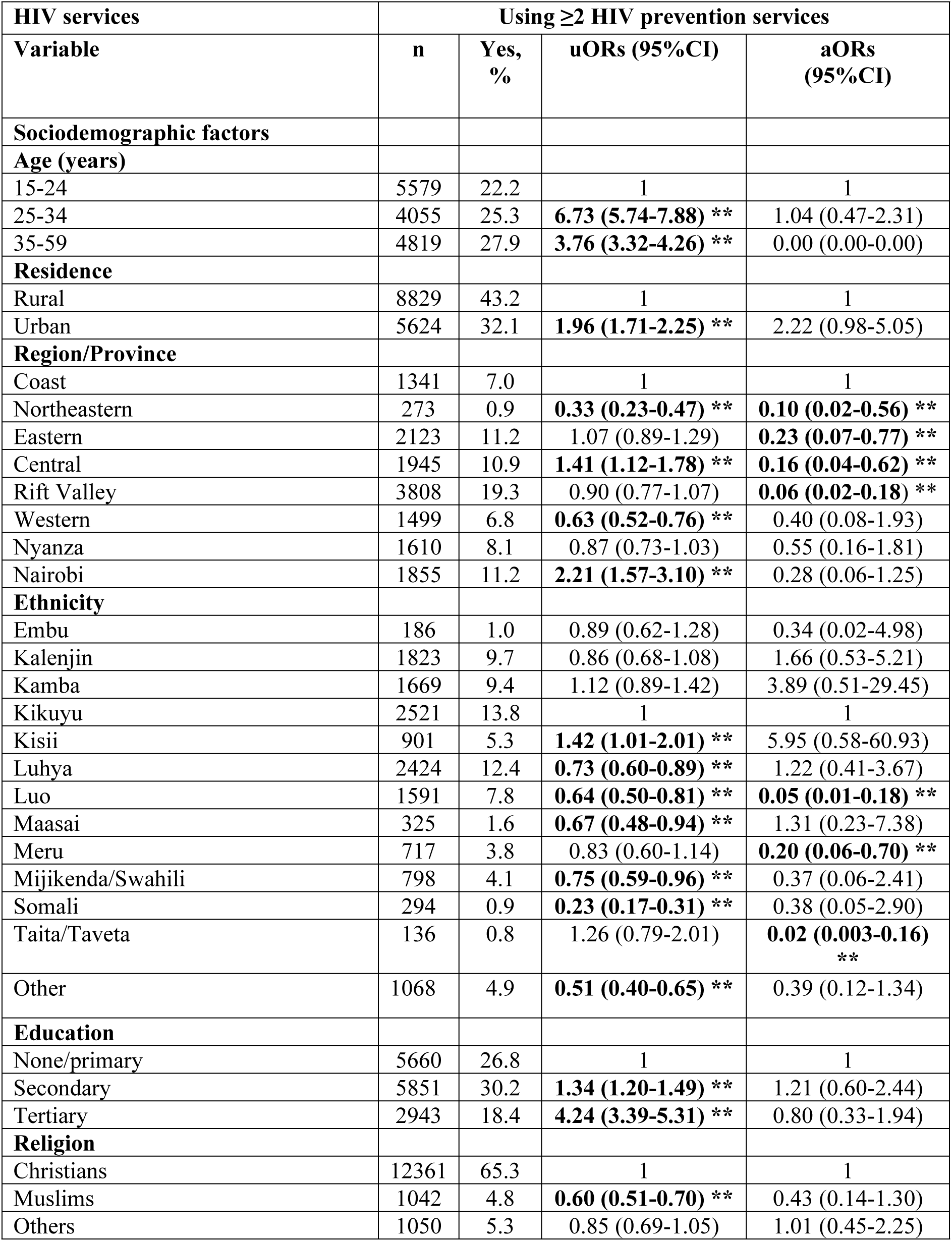
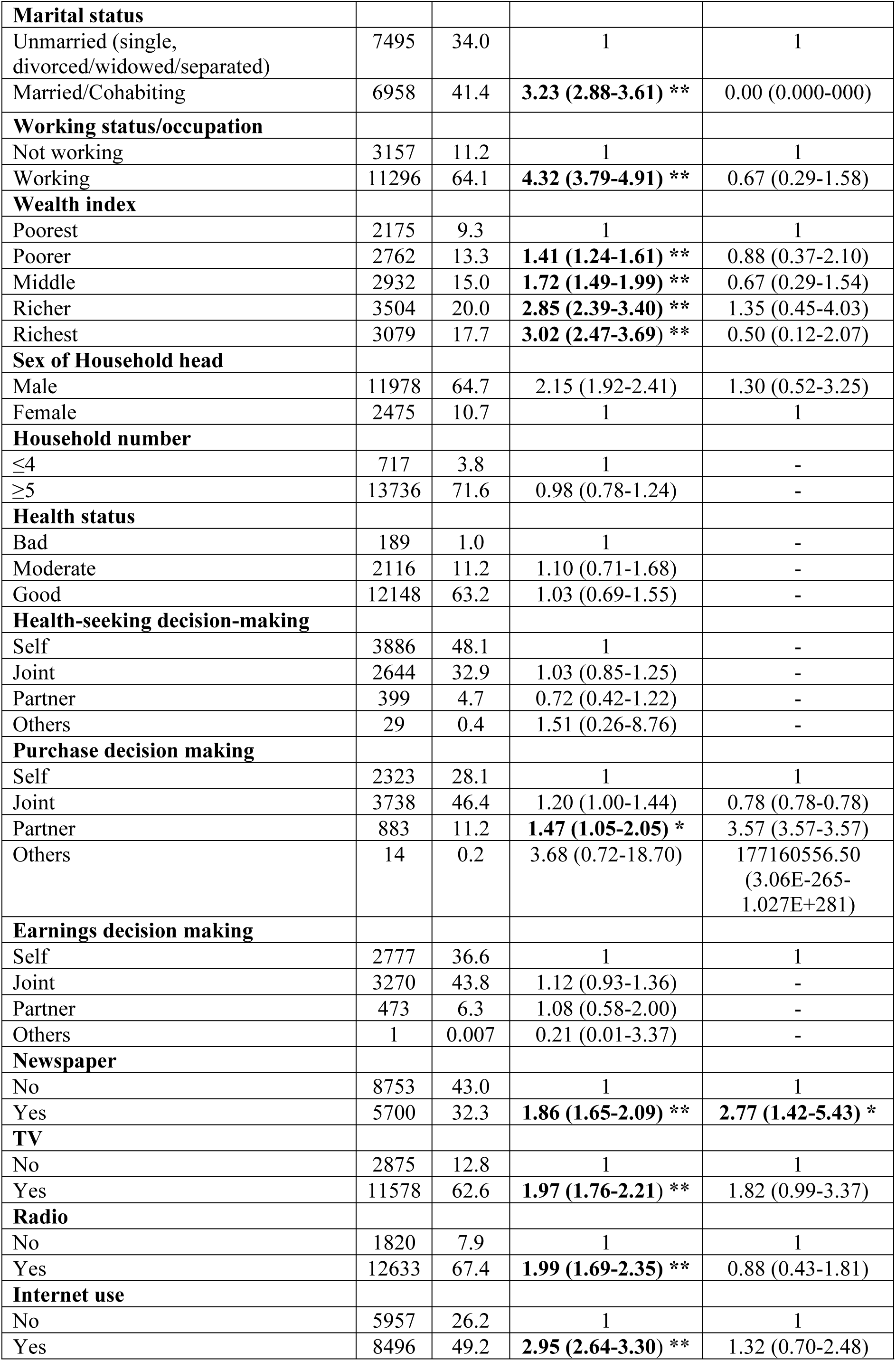
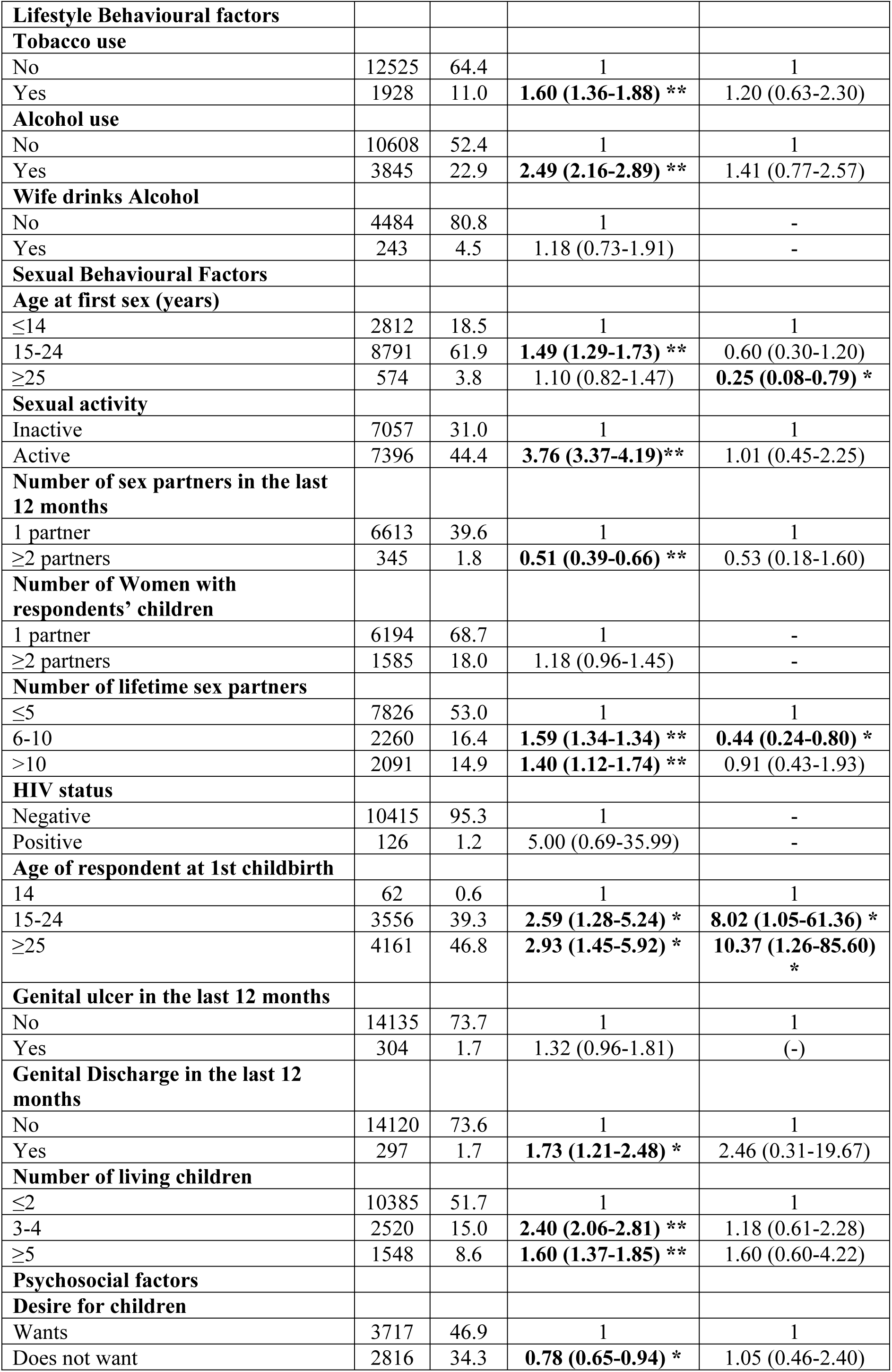
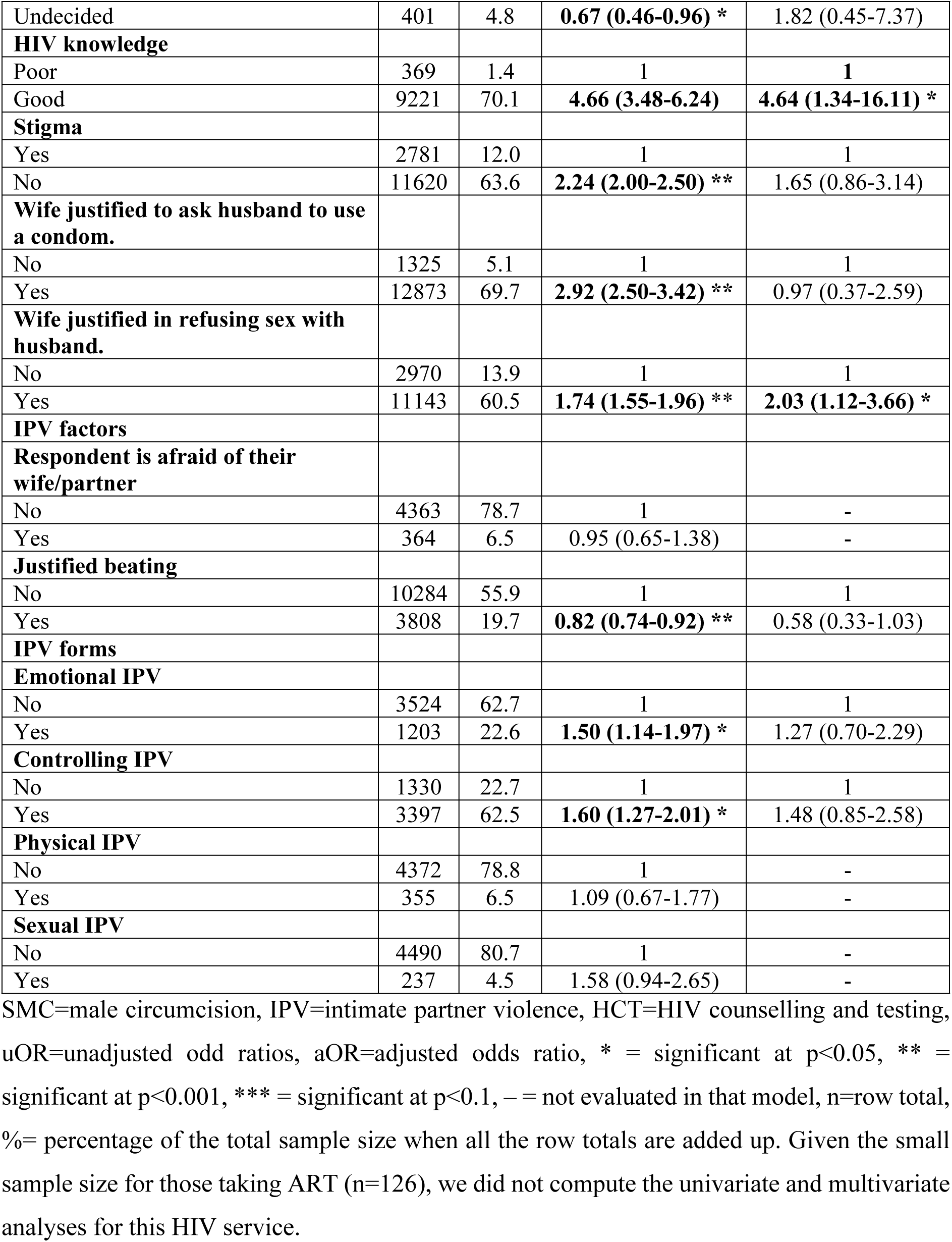
Factors associated with using multiple HIV prevention methods among men in Kenya

| HIV services | Using $\geq 2$ HIV prevention services | | | |
| --- | --- | --- | --- | --- |
| Variable | n | Yes, % | uORs (95%CI) | aORs (95%CI) |
| **Sociodemographic factors** |  |  |  |  |
| **Age (years)** |  |  |  |  |
| 15-24 | 5579 | 22.2 | 1 | 1 |
| 25-34 | 4055 | 25.3 | **6.73 (5.74-7.88) \*\*** | 1.04 (0.47-2.31) |
| 35-59 | 4819 | 27.9 | **3.76 (3.32-4.26) \*\*** | 0.00 (0.00-0.00) |
| **Residence** |  |  |  |  |
| Rural | 8829 | 43.2 | 1 | 1 |
| Urban | 5624 | 32.1 | **1.96 (1.71-2.25) \*\*** | 2.22 (0.98-5.05) |
| **Region/Province** |  |  |  |  |
| Coast | 1341 | 7.0 | 1 | 1 |
| Northeastern | 273 | 0.9 | **0.33 (0.23-0.47) \*\*** | **0.10 (0.02-0.56) \*\*** |
| Eastern | 2123 | 11.2 | 1.07 (0.89-1.29) | **0.23 (0.07-0.77) \*\*** |
| Central | 1945 | 10.9 | **1.41 (1.12-1.78) \*\*** | **0.16 (0.04-0.62) \*\*** |
| Rift Valley | 3808 | 19.3 | 0.90 (0.77-1.07) | **0.06 (0.02-0.18) \*\*** |
| Western | 1499 | 6.8 | **0.63 (0.52-0.76) \*\*** | 0.40 (0.08-1.93) |
| Nyanza | 1610 | 8.1 | 0.87 (0.73-1.03) | 0.55 (0.16-1.81) |
| Nairobi | 1855 | 11.2 | **2.21 (1.57-3.10) \*\*** | 0.28 (0.06-1.25) |
| **Ethnicity** |  |  |  |  |
| Embu | 186 | 1.0 | 0.89 (0.62-1.28) | 0.34 (0.02-4.98) |
| Kalenjin | 1823 | 9.7 | 0.86 (0.68-1.08) | 1.66 (0.53-5.21) |
| Kamba | 1669 | 9.4 | 1.12 (0.89-1.42) | 3.89 (0.51-29.45) |
| Kikuyu | 2521 | 13.8 | 1 | 1 |
| Kisii | 901 | 5.3 | **1.42 (1.01-2.01) \*\*** | 5.95 (0.58-60.93) |
| Luhya | 2424 | 12.4 | **0.73 (0.60-0.89) \*\*** | 1.22 (0.41-3.67) |
| Luo | 1591 | 7.8 | **0.64 (0.50-0.81) \*\*** | **0.05 (0.01-0.18) \*\*** |
| Maasai | 325 | 1.6 | **0.67 (0.48-0.94) \*\*** | 1.31 (0.23-7.38) |
| Meru | 717 | 3.8 | 0.83 (0.60-1.14) | **0.20 (0.06-0.70) \*\*** |
| Mijikenda/Swahili | 798 | 4.1 | **0.75 (0.59-0.96) \*\*** | 0.37 (0.06-2.41) |
| Somali | 294 | 0.9 | **0.23 (0.17-0.31) \*\*** | 0.38 (0.05-2.90) |
| Taita/Taveta | 136 | 0.8 | 1.26 (0.79-2.01) | **0.02 (0.003-0.16) \*\*** |
| Other | 1068 | 4.9 | **0.51 (0.40-0.65) \*\*** | 0.39 (0.12-1.34) |
| **Education** |  |  |  |  |
| None/primary | 5660 | 26.8 | 1 | 1 |
| Secondary | 5851 | 30.2 | **1.34 (1.20-1.49) \*\*** | 1.21 (0.60-2.44) |
| Tertiary | 2943 | 18.4 | **4.24 (3.39-5.31) \*\*** | 0.80 (0.33-1.94) |
| **Religion** |  |  |  |  |
| Christians | 12361 | 65.3 | 1 | 1 |
| Muslims | 1042 | 4.8 | **0.60 (0.51-0.70) \*\*** | 0.43 (0.14-1.30) |
| Others | 1050 | 5.3 | 0.85 (0.69-1.05) | 1.01 (0.45-2.25) |
| **Marital status** |  |  |  |  |
| Unmarried (single, divorced/widowed/separated) | 7495 | 34.0 | 1 | 1 |
| Married/Cohabiting | 6958 | 41.4 | **3.23 (2.88-3.61) \*\*** | 0.00 (0.000-0.000) |
| **Working status/occupation** |  |  |  |  |
| Not working | 3157 | 11.2 | 1 | 1 |
| Working | 11296 | 64.1 | **4.32 (3.79-4.91) \*\*** | 0.67 (0.29-1.58) |
| **Wealth index** |  |  |  |  |
| Poorest | 2175 | 9.3 | 1 | 1 |
| Poorer | 2762 | 13.3 | **1.41 (1.24-1.61) \*\*** | 0.88 (0.37-2.10) |
| Middle | 2932 | 15.0 | **1.72 (1.49-1.99) \*\*** | 0.67 (0.29-1.54) |
| Richer | 3504 | 20.0 | **2.85 (2.39-3.40) \*\*** | 1.35 (0.45-4.03) |
| Richest | 3079 | 17.7 | **3.02 (2.47-3.69) \*\*** | 0.50 (0.12-2.07) |
| **Sex of Household head** |  |  |  |  |
| Male | 11978 | 64.7 | 2.15 (1.92-2.41) | 1.30 (0.52-3.25) |
| Female | 2475 | 10.7 | 1 | 1 |
| **Household number** |  |  |  |  |
| ≤4 | 717 | 3.8 | 1 | - |
| ≥5 | 13736 | 71.6 | 0.98 (0.78-1.24) | - |
| **Health status** |  |  |  |  |
| Bad | 189 | 1.0 | 1 | - |
| Moderate | 2116 | 11.2 | 1.10 (0.71-1.68) | - |
| Good | 12148 | 63.2 | 1.03 (0.69-1.55) | - |
| **Health-seeking decision-making** |  |  |  |  |
| Self | 3886 | 48.1 | 1 | - |
| Joint | 2644 | 32.9 | 1.03 (0.85-1.25) | - |
| Partner | 399 | 4.7 | 0.72 (0.42-1.22) | - |
| Others | 29 | 0.4 | 1.51 (0.26-8.76) | - |
| **Purchase decision making** |  |  |  |  |
| Self | 2323 | 28.1 | 1 | 1 |
| Joint | 3738 | 46.4 | 1.20 (1.00-1.44) | 0.78 (0.78-0.78) |
| Partner | 883 | 11.2 | **1.47 (1.05-2.05) \*** | 3.57 (3.57-3.57) |
| Others | 14 | 0.2 | 3.68 (0.72-18.70) | 177160556.50<br>(3.06E-265-1.027E+281) |
| **Earnings decision making** |  |  |  |  |
| Self | 2777 | 36.6 | 1 | 1 |
| Joint | 3270 | 43.8 | 1.12 (0.93-1.36) | - |
| Partner | 473 | 6.3 | 1.08 (0.58-2.00) | - |
| Others | 1 | 0.007 | 0.21 (0.01-3.37) | - |
| **Newspaper** |  |  |  |  |
| No | 8753 | 43.0 | 1 | 1 |
| Yes | 5700 | 32.3 | **1.86 (1.65-2.09) \*\*** | **2.77 (1.42-5.43) \*** |
| **TV** |  |  |  |  |
| No | 2875 | 12.8 | 1 | 1 |
| Yes | 11578 | 62.6 | **1.97 (1.76-2.21) \*\*** | 1.82 (0.99-3.37) |
| **Radio** |  |  |  |  |
| No | 1820 | 7.9 | 1 | 1 |
| Yes | 12633 | 67.4 | **1.99 (1.69-2.35) \*\*** | 0.88 (0.43-1.81) |
| **Internet use** |  |  |  |  |
| No | 5957 | 26.2 | 1 | 1 |
| Yes | 8496 | 49.2 | **2.95 (2.64-3.30) \*\*** | 1.32 (0.70-2.48) |
| **Lifestyle Behavioural factors** |  |  |  |  |
| **Tobacco use** |  |  |  |  |
| No | 12525 | 64.4 | 1 | 1 |
| Yes | 1928 | 11.0 | **1.60 (1.36-1.88) \*\*** | 1.20 (0.63-2.30) |
| **Alcohol use** |  |  |  |  |
| No | 10608 | 52.4 | 1 | 1 |
| Yes | 3845 | 22.9 | **2.49 (2.16-2.89) \*\*** | 1.41 (0.77-2.57) |
| **Wife drinks Alcohol** |  |  |  |  |
| No | 4484 | 80.8 | 1 | - |
| Yes | 243 | 4.5 | 1.18 (0.73-1.91) | - |
| **Sexual Behavioural Factors** |  |  |  |  |
| **Age at first sex (years)** |  |  |  |  |
| ≤14 | 2812 | 18.5 | 1 | 1 |
| 15-24 | 8791 | 61.9 | **1.49 (1.29-1.73) \*\*** | 0.60 (0.30-1.20) |
| ≥25 | 574 | 3.8 | 1.10 (0.82-1.47) | **0.25 (0.08-0.79) \*** |
| **Sexual activity** |  |  |  |  |
| Inactive | 7057 | 31.0 | 1 | 1 |
| Active | 7396 | 44.4 | **3.76 (3.37-4.19)\*\*** | 1.01 (0.45-2.25) |
| **Number of sex partners in the last 12 months** |  |  |  |  |
| 1 partner | 6613 | 39.6 | 1 | 1 |
| ≥2 partners | 345 | 1.8 | **0.51 (0.39-0.66) \*\*** | 0.53 (0.18-1.60) |
| **Number of Women with respondents' children** |  |  |  |  |
| 1 partner | 6194 | 68.7 | 1 | - |
| ≥2 partners | 1585 | 18.0 | 1.18 (0.96-1.45) | - |
| **Number of lifetime sex partners** |  |  |  |  |
| ≤5 | 7826 | 53.0 | 1 | 1 |
| 6-10 | 2260 | 16.4 | **1.59 (1.34-1.34) \*\*** | **0.44 (0.24-0.80) \*** |
| >10 | 2091 | 14.9 | **1.40 (1.12-1.74) \*\*** | 0.91 (0.43-1.93) |
| **HIV status** |  |  |  |  |
| Negative | 10415 | 95.3 | 1 | - |
| Positive | 126 | 1.2 | 5.00 (0.69-35.99) | - |
| **Age of respondent at 1st childbirth** |  |  |  |  |
| 14 | 62 | 0.6 | 1 | 1 |
| 15-24 | 3556 | 39.3 | **2.59 (1.28-5.24) \*** | **8.02 (1.05-61.36) \*** |
| ≥25 | 4161 | 46.8 | **2.93 (1.45-5.92) \*** | **10.37 (1.26-85.60) \*** |
| **Genital ulcer in the last 12 months** |  |  |  |  |
| No | 14135 | 73.7 | 1 | 1 |
| Yes | 304 | 1.7 | 1.32 (0.96-1.81) | (-) |
| **Genital Discharge in the last 12 months** |  |  |  |  |
| No | 14120 | 73.6 | 1 | 1 |
| Yes | 297 | 1.7 | **1.73 (1.21-2.48) \*** | 2.46 (0.31-19.67) |
| **Number of living children** |  |  |  |  |
| ≤2 | 10385 | 51.7 | 1 | 1 |
| 3-4 | 2520 | 15.0 | **2.40 (2.06-2.81) \*\*** | 1.18 (0.61-2.28) |
| ≥5 | 1548 | 8.6 | **1.60 (1.37-1.85) \*\*** | 1.60 (0.60-4.22) |
| **Psychosocial factors** |  |  |  |  |
| **Desire for children** |  |  |  |  |
| Wants | 3717 | 46.9 | 1 | 1 |
| Does not want | 2816 | 34.3 | **0.78 (0.65-0.94) \*** | 1.05 (0.46-2.40) |
| Undecided | 401 | 4.8 | **0.67 (0.46-0.96) \*** | 1.82 (0.45-7.37) |
| **HIV knowledge** |  |  |  |  |
| Poor | 369 | 1.4 | 1 | 1 |
| Good | 9221 | 70.1 | **4.66 (3.48-6.24)** | **4.64 (1.34-16.11) \*** |
| **Stigma** |  |  |  |  |
| Yes | 2781 | 12.0 | 1 | 1 |
| No | 11620 | 63.6 | **2.24 (2.00-2.50) \*\*** | 1.65 (0.86-3.14) |
| **Wife justified to ask husband to use a condom.** |  |  |  |  |
| No | 1325 | 5.1 | 1 | 1 |
| Yes | 12873 | 69.7 | **2.92 (2.50-3.42) \*\*** | 0.97 (0.37-2.59) |
| **Wife justified in refusing sex with husband.** |  |  |  |  |
| No | 2970 | 13.9 | 1 | 1 |
| Yes | 11143 | 60.5 | **1.74 (1.55-1.96) \*\*** | **2.03 (1.12-3.66) \*** |
| **IPV factors** |  |  |  |  |
| **Respondent is afraid of their wife/partner** |  |  |  |  |
| No | 4363 | 78.7 | 1 | - |
| Yes | 364 | 6.5 | 0.95 (0.65-1.38) | - |
| **Justified beating** |  |  |  |  |
| No | 10284 | 55.9 | 1 | 1 |
| Yes | 3808 | 19.7 | **0.82 (0.74-0.92) \*\*** | 0.58 (0.33-1.03) |
| **IPV forms** |  |  |  |  |
| **Emotional IPV** |  |  |  |  |
| No | 3524 | 62.7 | 1 | 1 |
| Yes | 1203 | 22.6 | **1.50 (1.14-1.97) \*** | 1.27 (0.70-2.29) |
| **Controlling IPV** |  |  |  |  |
| No | 1330 | 22.7 | 1 | 1 |
| Yes | 3397 | 62.5 | **1.60 (1.27-2.01) \*** | 1.48 (0.85-2.58) |
| **Physical IPV** |  |  |  |  |
| No | 4372 | 78.8 | 1 | - |
| Yes | 355 | 6.5 | 1.09 (0.67-1.77) | - |
| **Sexual IPV** |  |  |  |  |
| No | 4490 | 80.7 | 1 | - |
| Yes | 237 | 4.5 | 1.58 (0.94-2.65) | - |

SMC=male circumcision, IPV=intimate partner violence, HCT=HIV counselling and testing, uOR=unadjusted odd ratios, aOR=adjusted odds ratio, \* = significant at $p<0.05$ , \*\* = significant at $p<0.001$ , \*\*\* = significant at $p<0.1$ , – = not evaluated in that model, n=row total, %= percentage of the total sample size when all the row totals are added up. Given the small sample size for those taking ART (n=126), we did not compute the univariate and multivariate analyses for this HIV service.

## Discussion

Suppressing the HIV scourge among men needs continuous and effective use of multiple HIV prevention services. Antecedent to this study, there was evidence on the use of more than one HIV prevention measure as a way to reduce HIV spread in risk groups. However, literature about the use of multiple HIV services among men in the general population, especially in Kenya, was limited. This study found that the majority (75.4%) had used multiple HIV services (>2), while 98% of the men in Kenya had used at least one of the four HIV prevention methods. Additionally, a relatively low proportion of men used multiple HIV services. For instance, 22.7% of the men had tested for HIV, were circumcised, and then used condoms, followed by men who had tested for HIV, and then used condoms (23.9%), and those who were circumcised and then used condoms (28.7%). These study findings contrast with the previous studies conducted in Nigeria and Uganda, which reported the prevalence of the use of multiple HIV preventive services among men to be 2.8% and 45% (11, 16). The reasons for the low utilisation in Nigeria when compared with Kenya and Uganda may include lack of awareness and inaccessibility to HIV services, and fear of the consequences in case one tests HIV positive (17, 18). The Kenyan government should strengthen and empower interventions such as free condom provision, free HIV testing services, and increased HIV awareness campaigns to ensure effective utilisation of multiple HIV services among men to prevent and reduce HIV transmission in the general population in Kenya In multivariable analysis, we found several factors associated with the use of multiple HIV services, such as HIV Knowledge, ethnicity, age at first sex, age at first child, number of sex partners, and access to media information. This study found that men with good knowledge of HIV were significantly more likely to use multiple HIV prevention methods compared with those with poor knowledge. These study findings concur with previous studies that demonstrated a strong positive correlation between HIV-related knowledge and the adoption of HIV preventive measures (19–21). Increased awareness enhances risk perception and empowers individuals to make informed decisions regarding HIV prevention strategies such as condom use, pre-exposure prophylaxis (PrEP), and voluntary medical male circumcision (VMMC). Thus, the stakeholders should strengthen and sustain awareness campaigns in the community so that men can gain a better understanding of the benefits of using HIV prevention services, thus increasing their uptake.

Our study findings further indicated that men who had their first sexual experience at an older age (≥25 years) were significantly less likely to use multiple HIV prevention methods. This finding may reflect the possibility that delayed sexual debut is often associated with a perception of lower risk and access to youth-friendly services, thereby reducing the motivation to engage in multiple prevention strategies (22). This concurs with a study that reported similar findings(22). On the contrary, men who had their first child at an older age (≥25 years) were more likely to use multiple HIV prevention methods. This suggests that late fatherhood may be associated with greater responsibility and health-conscious behaviours, leading to the adoption of diverse HIV prevention measures (22). We recommend continuous education programmes about HIV prevention among married men (or men with later sexual debut) through targeted peer-to-peer and digital approaches as well as additional qualitative studies to comprehensively comprehend why men at an older age (≥25) were less likely to use multiple HIV services.

Our study also found that men with a high number of lifetime sexual partners (6-10 partners) were less likely to use multiple HIV prevention methods compared with those with fewer partners (≤5). This finding corroborates existing research suggesting that individuals with higher sexual exposure may paradoxically engage in fewer preventive measures, possibly due to risk normalization or behavioral disinhibition (23–25). Targeted interventions involving continuous awareness campaigns and behaviour change interventions that emphasize the HIV risk and benefits of using multiple HIV prevention services are needed to address this gap among sexually active men with multiple sex partners.

In this study, we found out that access to information was a crucial determinant of HIV prevention behaviours. Men with access to newspapers were significantly more likely to use multiple HIV prevention methods. This study concurs with the findings of a study that reported that men in South Africa who often had access to televisions and newspapers frequently went for HIV testing and used condoms regularly (26). This finding supports the role of media and information dissemination in influencing health behaviours (27). Ensuring widespread access to accurate HIV information through various media platforms can be a vital strategy in improving HIV prevention efforts. This study emphasizes the need for targeted HIV prevention programs that enhance knowledge dissemination, address behavioral risks, and promote access to reliable information. Future interventions should focus on leveraging mass media channels, promoting comprehensive sexual education, and tailoring prevention strategies to men in high-risk populations.

Across the tribes of Kenya, Men from some tribes of Kenya had lower odds of using multiple HIV testing services when compared with the Kikuyu. This concurs with the previous study conducted in Kenya among the general population, which reported that some tribes were less involved in HIV prevention programs (28). One possible explanation for these findings could be that there exist negative cultural and religious beliefs on some HIV prevention methods that concurrently serve as family planning methods, such as condom use and circumcision (28). Tribes espousing such negative beliefs often engage in unprotected sex and other risky sexual behaviours. Therefore, stakeholders need to engage cultural leaders in HIV awareness and promotion of the use of HIV prevention services to curb the HIV spread in Kenya.

We found that men who believed it was justified for a woman to refuse sex with her partner were twice as likely to use multiple HIV services. This concurs with a study in Malawi, which found that men with gender-equitable attitudes were more likely to engage in HIV testing and prevention behaviors (29) This association may reflect a more progressive or gender equitable attitude, which is often linked with better health-seeking behaviors, including HIV testing and treatment (30). These men may be more open to communication about sexual health and perceive fewer social barriers to using HIV services. In contrast, our study differs from one study conducted in rural Uganda, which suggests that despite gender-equitable attitudes, structural barriers still limit HIV services uptake (31). Therefore, stakeholders, including the government of Kenya, should incorporate gender-transformative messaging in HIV outreach programs and mass media to promote HIV service use among men.

Finally, we found that Men from these regions some regions of Kenya were less likely to use multiple HIV services compared with those from the coastal region. The Northeastern region of Kenya showed extremely low odds, potentially due to cultural norms, stigma, limited infrastructure, insecurity, and a predominantly pastoralist population that may face mobility-related barriers. This study’s findings also concur with a study that found regional disparities in HIV service utilization in Kenya, with urban areas outperforming rural and arid regions. (32) The Kenya National AIDS Control Council (NACC) report (2020) also identified the Northeastern region as underperforming in HIV service coverage, partly due to poor road infrastructure and health facility density in the Northeastern and Rift Valley, which limited physical access. This may also be influenced by taboos around discussing sexual health, which may limit HIV service uptake. In contrast, some programmatic data from NGOs working in Eastern and Rift Valley regions of Kenya suggested that targeted mobile outreach programs improved uptake in hard-to-reach areas (33). However, such success was highly dependent on consistent donor support and community engagement. We recommend that the government of Kenya implement region-specific interventions, including mobile clinics and culturally sensitive health promotion, strengthening community health systems in underserved regions, and collaborating with local leaders and influencers to reduce stigma and improve awareness of HIV services.

## Strengths and limitations of the study

This study investigated the use of multiple HIV prevention services among men in Kenya, unlike previous studies, which focused on a single use of HIV prevention services. Furthermore, the study utilized data from the 2022 Kenya Demographic and Health Survey (KDHS), a nationally representative survey that enhances the generalizability of the findings to the male population across Kenya. The use of a large and diverse sample provides robust statistical power and reduces the likelihood of biased estimates, enabling more reliable analysis of factors associated with the use of multiple HIV prevention services. By examining various demographic, cultural, and behavioral factors, the study provides a holistic understanding of the determinants influencing multiple HIV prevention service uptake among men.

However, because the study used self-reported measures, when evaluating concepts such as sexual behaviours, it may introduce social desirability, potentially affecting the accuracy of the findings. While the study explores cultural factors, the KDHS might not fully capture nuanced community-specific beliefs and practices that influence HIV prevention behaviours. In addition, the study does not provide information on changes in HIV prevention behaviour over time, which could help assess the impact of past or ongoing interventions. Lastly, some study results had wider confidence intervals, making some of our findings imprecise in predicting the use of multiple HIV Services. Thus, more studies with DHS are needed in Kenya and other countries.

## Conclusions

Suppressing the HIV epidemic among men requires continuous and effective utilization of multiple HIV prevention services. This study highlights that a significant proportion of men in Kenya (75.4%) used multiple HIV prevention methods, a promising finding compared to lower utilization rates reported in other countries, such as Nigeria and Uganda.

The study underscores the importance of multilevel strategies in influencing the use of multiple HIV prevention services, such as knowledge on use of HIV self-testing, lifetime partners, region, age, access to media, and tribe. Stakeholders need to involve and incorporate cultural and religious leaders to raise awareness about the benefits of prevention services and ensure their accessibility. There is a need to incorporate gender-transformative messaging in HIV outreach programs to promote HIV service use among men. The government of Kenya should implement region-specific interventions, including mobile clinics and culturally sensitive health promotion, to strengthen community health systems in underserved regions.

## Data Availability

Third-party data was obtained for this study from The DHS Program (https://dhsprogram.com/). Data may be requested from the DHS Program after creating an account and submitting a concept note. More access information can be found on the DHS Program website (https://dhsprogram.com/data/Access-Instructions.cfm). The data set is openly available upon permission from the MEASURE DHS website (https://www.dhsprogram.com/data/available-datasets.cfm). The authors confirm that interested researchers would be able to access these data in the same manner as the authors. The authors also confirm that they had no special access privileges that others would not have

## Abbreviations

PrEP: Pre-Exposure Prophylaxis
WHO: World Health Organization
VMMC: Voluntary Medical Male Circumcision
HTC: HIV Testing and Counselling
EMTC: Elimination of Mother-to-Child Transmission
ART: Antiretroviral Therapy
SMC: Safe male circumcision
EA: Enumeration area
FBO: Faith-based organization
aOR: Adjusted Odds Ratio
KDHS: Kenya Demographic Health Survey
CI: Confidence Interval
DHS: Demographic Health Survey
VIF: Variance Inflation Factor
uOR: Unadjusted Odds Ratio
OR: Odds Ratio
SPSS: Statistical Package for Social Science
NGO: Non-government organization

## DECLARATIONS

### Acknowledgments

We thank the DHS program for making the data available for this study.

